# Long-Term Outcomes of Successful CTO PCI Using Antegrade and Retrograde Dissection and Re-entry Technique

**DOI:** 10.1101/2025.09.10.25335543

**Authors:** Gao Haokao, Yu Tiantong, Wang Huan, Cai Yue, Wang Mian, Dou Kefei, Zhao Lin, Jin Zening, Hou Yinglong, Chen Youhu, Lei Xiaolin, Lian Kun, Chen Genrui, Li Chengxiang

## Abstract

**Background:** There are always concerns regarding the durability of subintimal stenting after contemporary chronic total occlusion (CTO) recanalization.

**Method:** This prospective, multicenter clinical trial aimed to compare extraplaque (EP) and intraplaque (IP) tracking for long-term clinical outcomes after CTO recanalization. After IVUS assessment of the wire crossing position, the patients were divided into two groups: (1) the EP group with subintimal length (SL) > 10 mm; and (2) the IP group with intraplaque or minor extraplaque (SL ≤ 10 mm). The primary endpoint was binary in-segment restenosis (ISR), and the secondary endpoint was the occurrence of major adverse cardiac events. Angiographic follow-up (FU) was scheduled at 13 months, and clinical FU was continued up to 3 years.

**Results:** A total of 257 successful CTO patients were enrolled. The mean CTO length in the EP group was 46.6 ± 16.1 mm, and the median extraplaque length was approximately 24.5 mm. The J-CTO score was higher (p<0.001). Angiographic FU at 13 months showed that both groups had similar cumulative rates of ISR (p = 0.704). In the 3-year clinical follow-up, the cumulative incidence of target vessel revascularization (TVR) in both groups and across different tracking techniques showed no significant difference (p > 0.05). Independent predictors of ISR were a total stent length >50 mm and a wire crossing time >2 hours, while multivessel lesions showed an independent association with 3-year TVR.

**Conclusion:** The specific long extraplaque tracking had comparable angiographic and long-term clinical outcomes to intraplaque tracking, regardless of the crossing technique used.

**Graphical Abstract:** 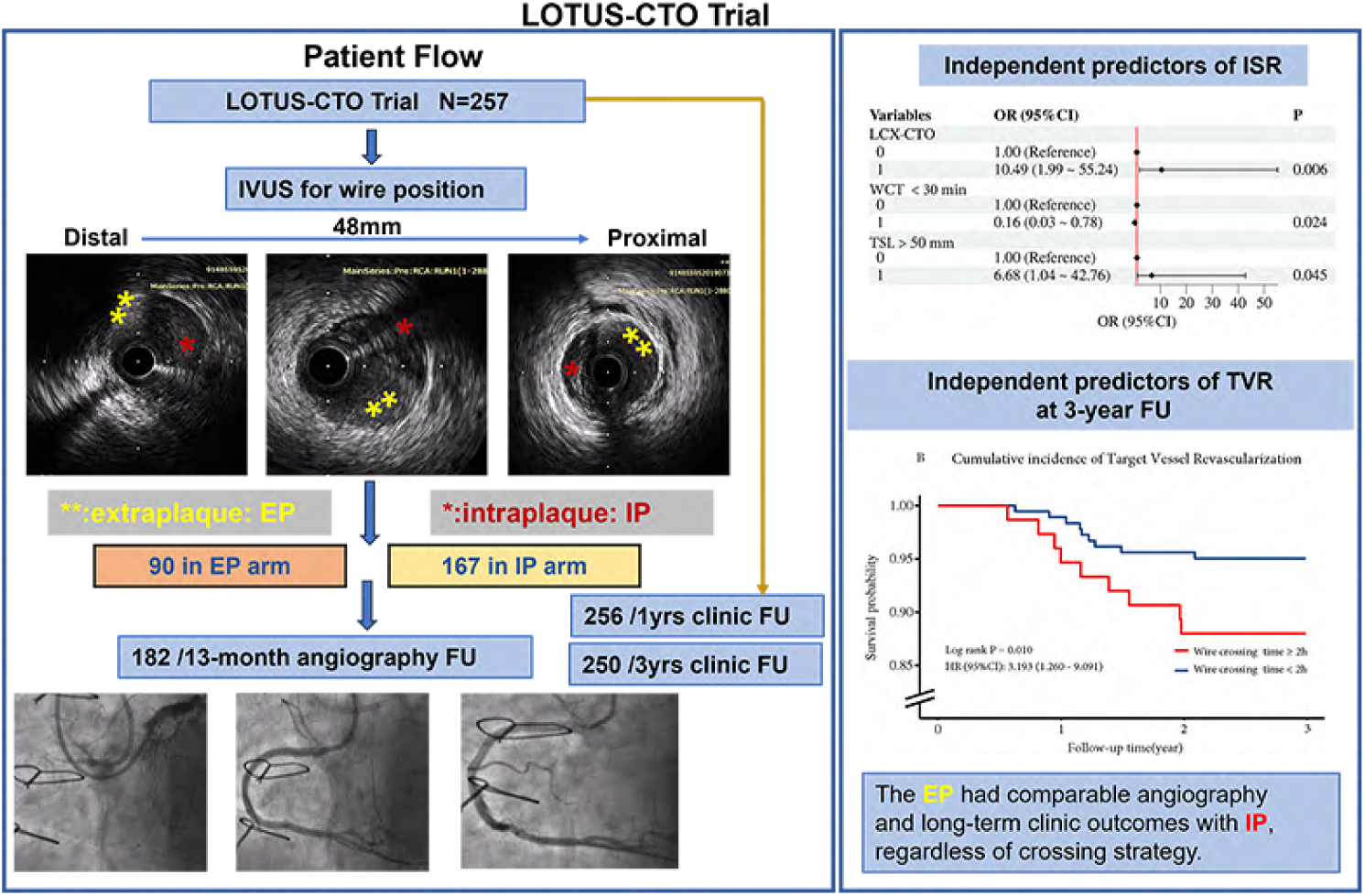

## Introduction

Chronic total occlusions (CTOs) represent the most challenging lesion subset to be treated with percutaneous coronary intervention (PCI) [1, 2]. A successful CTO PCI has been associated with improved clinical outcomes compared to failed procedures [3, 4]. The hybrid algorithm, particularly the development of dissection and re-entry techniques (DARTs), has improved success rates [5–7] accompanied by lower complication rates [8, 9]. In dedicated CTO-PCI centers, success rates of CTO-PCI have been reported to be as high as 95% [10].

However, there is an ongoing controversy regarding the relative merits of antegrade and retrograde wire escalation (AWE/RWE) versus dissection and reentry techniques (ADR/ RDR). Several studies have shown that extraplaque (EP) tracking does not adversely affect clinical outcomes at mid-term follow-up (FU) [11–19]. However, other individual studies and meta-analyses have shown an increased risk associated with EP tracking [20–23]. The inconsistencies among these studies need to be further clarified. As reported, multiple studies have assessed guidewire position based on technique tracking rather than systematic evaluation with intravascular ultrasound (IVUS) immediately after CTO crossing [12–15, 18, 22], which is to a certain extent imprecise. In a meta-analysis, the actual wire position differs from the expected position in approximately 10% of antegrade cases and 15%–30% of retrograde cases [23]. Meanwhile, a short subintimal track could have different clinical implications compared to a long track [24]; however, no study has addressed this specific question so far. Finally, systematic angiographic follow-up with a midterm evaluation was conducted in only a few studies [11, 15–18].

Therefore, the aim of this prospective study was to clarify the angiographic and long-term clinical outcomes of EP tracking following IVUS assessment of guidewire position after the application of contemporary CTO recanalization techniques.

## METHODS

### Patient Population and Trial Design

This prospective, multi-center registry trial (LOTUS CTO, Clinical Trial ID: NCT03769038) enrolled patients who underwent technically successful CTO-PCI (one CTO per patient) at one of the six participating centers in China.

The CTO treatment strategy was selected by operators at their discretion based on any of the available crossing algorithms. After completing a technically successful CTO-PCI, IVUS was used to evaluate the guide wire (GW) crossing position, and lesions were classified into two groups: (1) the Extraplaque (EP) group with subintimal length (SL) > 10 mm; and (2) the Intraplaque (IP) group, defined as true-to-true lumen or minor extraplaque with SL ≤ 10 mm. IVUS examinations were performed to optimize the implanted everolimus-eluting cobalt chromium stents (XIENCE PRIME™/XIENCE V, Abbott Vascular). Clinical, procedural, and imaging data, as well as follow-up data, were collected according to a standardized protocol and entered into a central electronic database.

### Study End Points and Definitions

The primary endpoint of the study was binary in-stent restenosis (ISR), defined as an angiographic diameter stenosis of 50% or more within the stented segment or within a 5 mm segment adjacent to the stent, based on QCA parameters obtained at 13 months. The secondary endpoint was the cumulative incidence of major adverse cardiac events (MACE), defined as a composite of all-cause death, cardiovascular death, myocardial infarction (MI), and target vessel revascularization (TVR). Additional endpoints included the percentage of LLL (an angiographic endpoint), and acute and late stent thrombosis in each recanalization strategy group. Surveillance angiography was scheduled 13 months after PCI. Clinical FU was conducted at 1 year and 3 years via phone interviews, review of hospital records, or outpatient visits.

## RESULTS

A total of 257 patients were enrolled between December 2018 and September 2021; these patients underwent successful CTO PCI procedures and were subsequently evaluated by IVUS to assess the GW position in the current study. Among them, 35% (90/257) were assigned to the EP group and 65% (167/257) to the IP group. The trial flow of the study is outlined in Figure 1.

**Figure 1.**
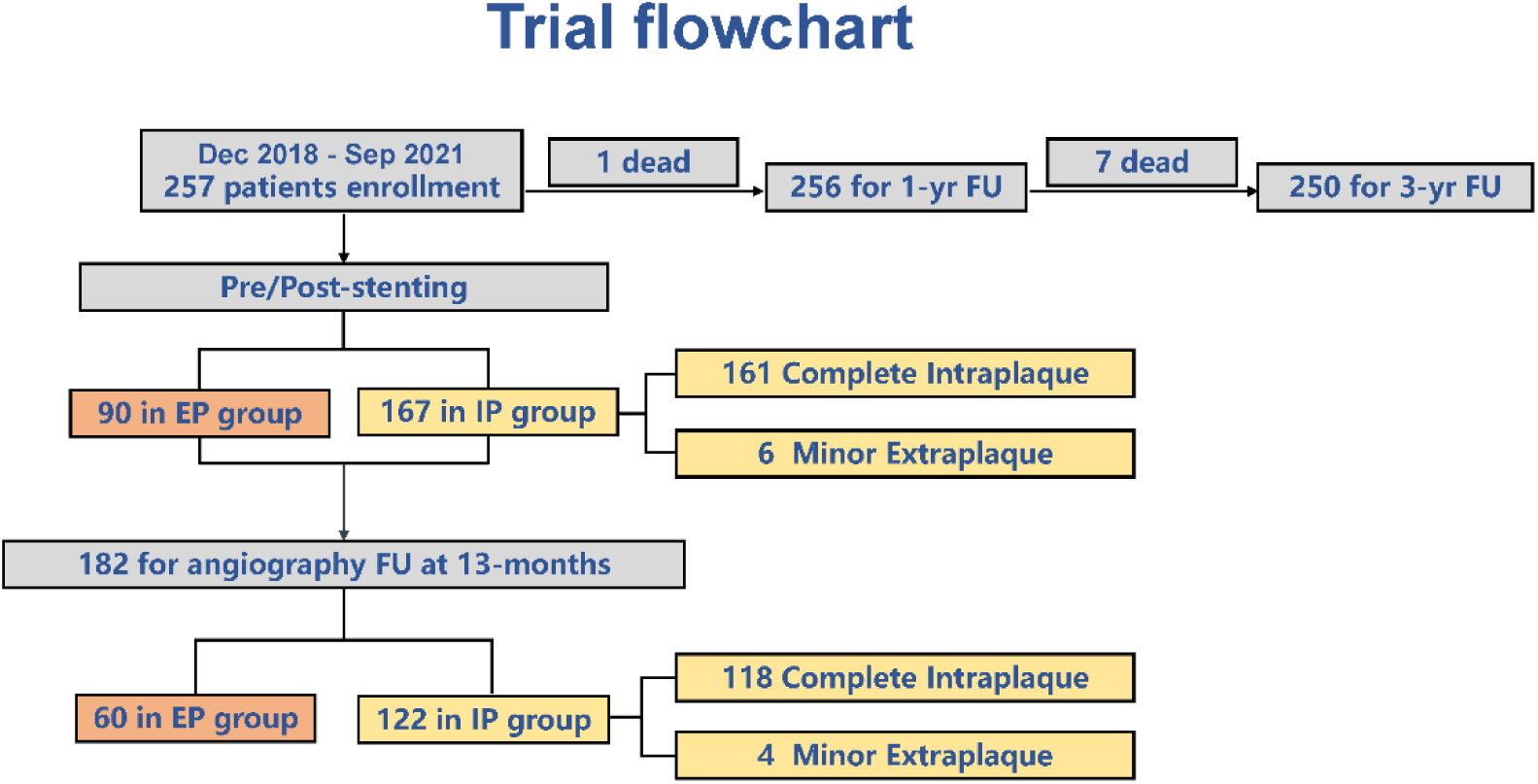
Flow Chart. EP: Extra-plaque; IP: Intraplaque; FU: Follow-Up. IVUS: Intravascular ultrasound;

### Baseline Demographics and Clinical Characteristics

As seen in Table 1, the mean age was 59.8 ± 10.5 years; 30.7% of the patients had a previous MI, 40.5% had a previous PCI, and 20.2% had undergone CABG surgery. There was a higher prevalence of previous PCI in the EP group than in the IP group (48.9% vs. 35.9%, *p* = 0.043). All other baseline demographics and clinical presentations were well balanced between the two groups. Additionally, the DART tracking group had higher LDL-c levels and a higher rate of hypertension compared to the no-DART group (all *p* < 0.05; see Table S1).

**Table 1.**
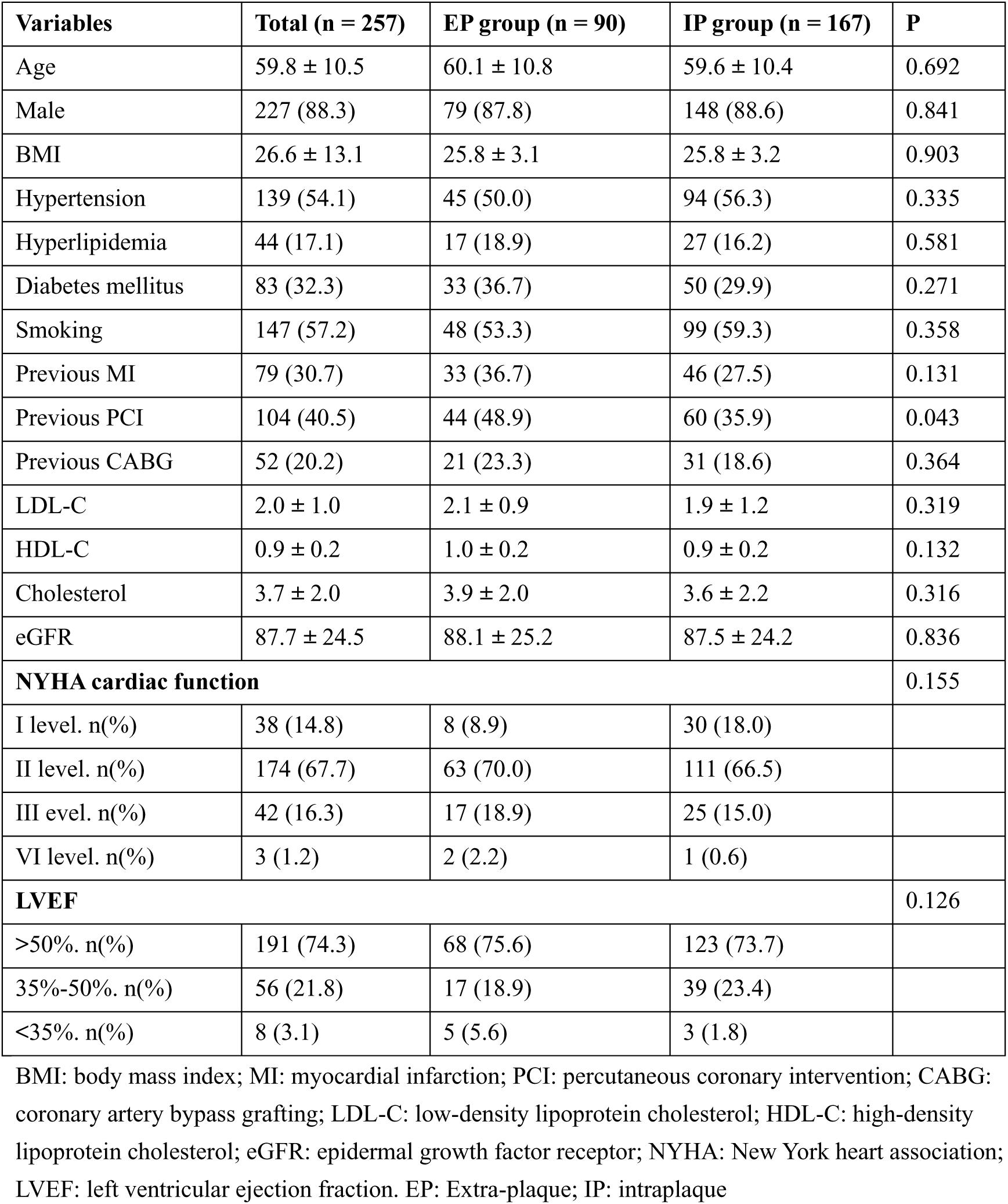
Baseline Clinical Characteristics in EP and IP groups.

### Angiography and Procedure Characteristics

As seen in Table 2, the right coronary artery (RCA) was the most common CTO target vessel (52.5%), followed by the left anterior descending artery (LAD) (40.5%). In the overall group, 95.7% of patients had a CTO length > 20 mm. The values of lesion characteristics were more complex without significant different between the two groups. The J-CTO score was high among all patients (2.28 ± 0.83). There was a statistically significant difference in the mean J-CTO score (2.54 ± 0.84 vs. 2.14 ± 0.80, *p* < 0.001) and in the proportion of a J-CTO score ≥3 (54.4% vs. 38.3%, *p* = 0.013) in the EP group compared with the IP group. There was no significant difference in the mean J-CTO score between the DART and the no-DART tracking groups (2.40 ± 0.88 vs. 2.24 ± 0.82, *p* = 0.182), as shown in Table S2.

**Table 2.**
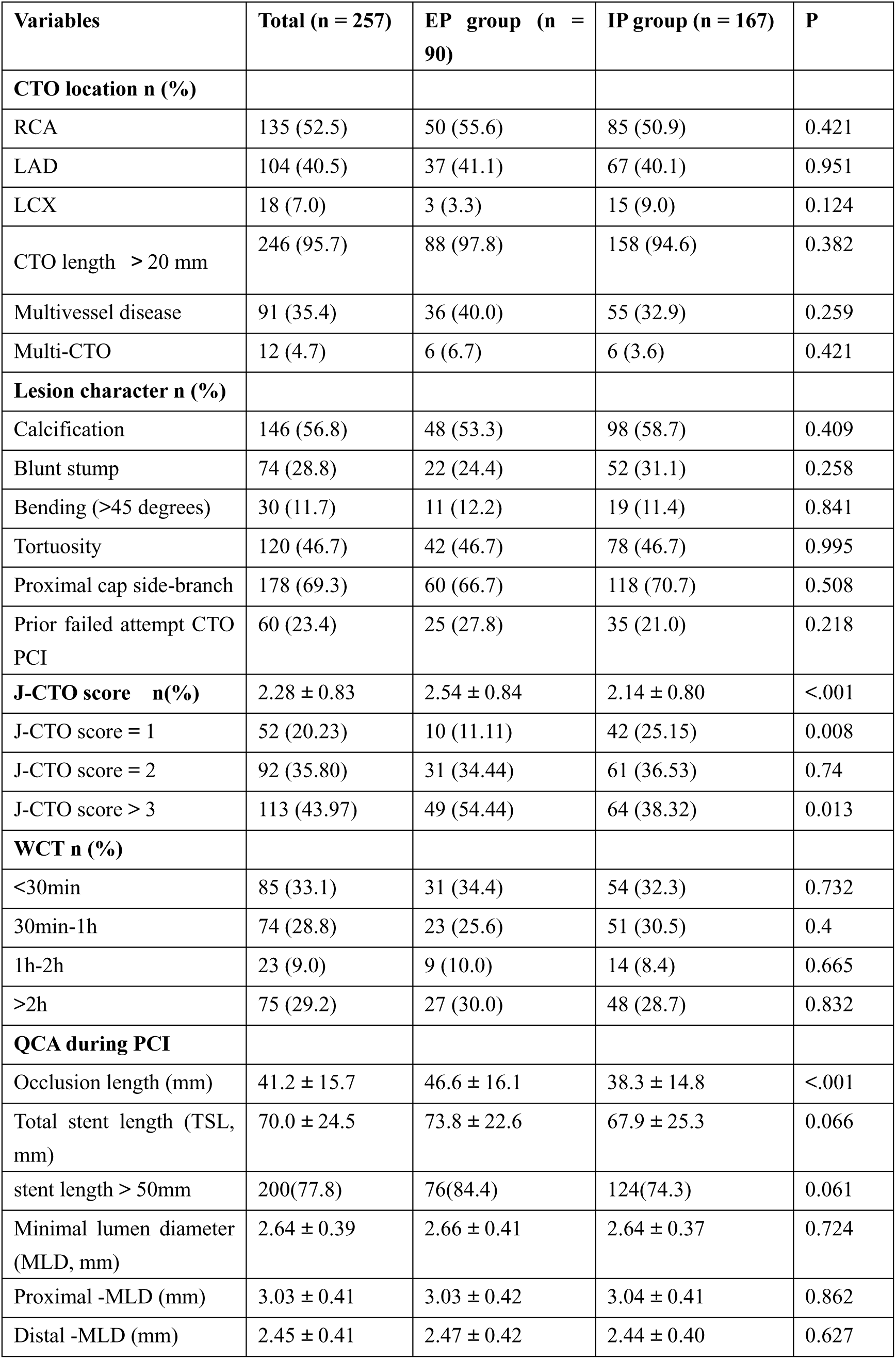
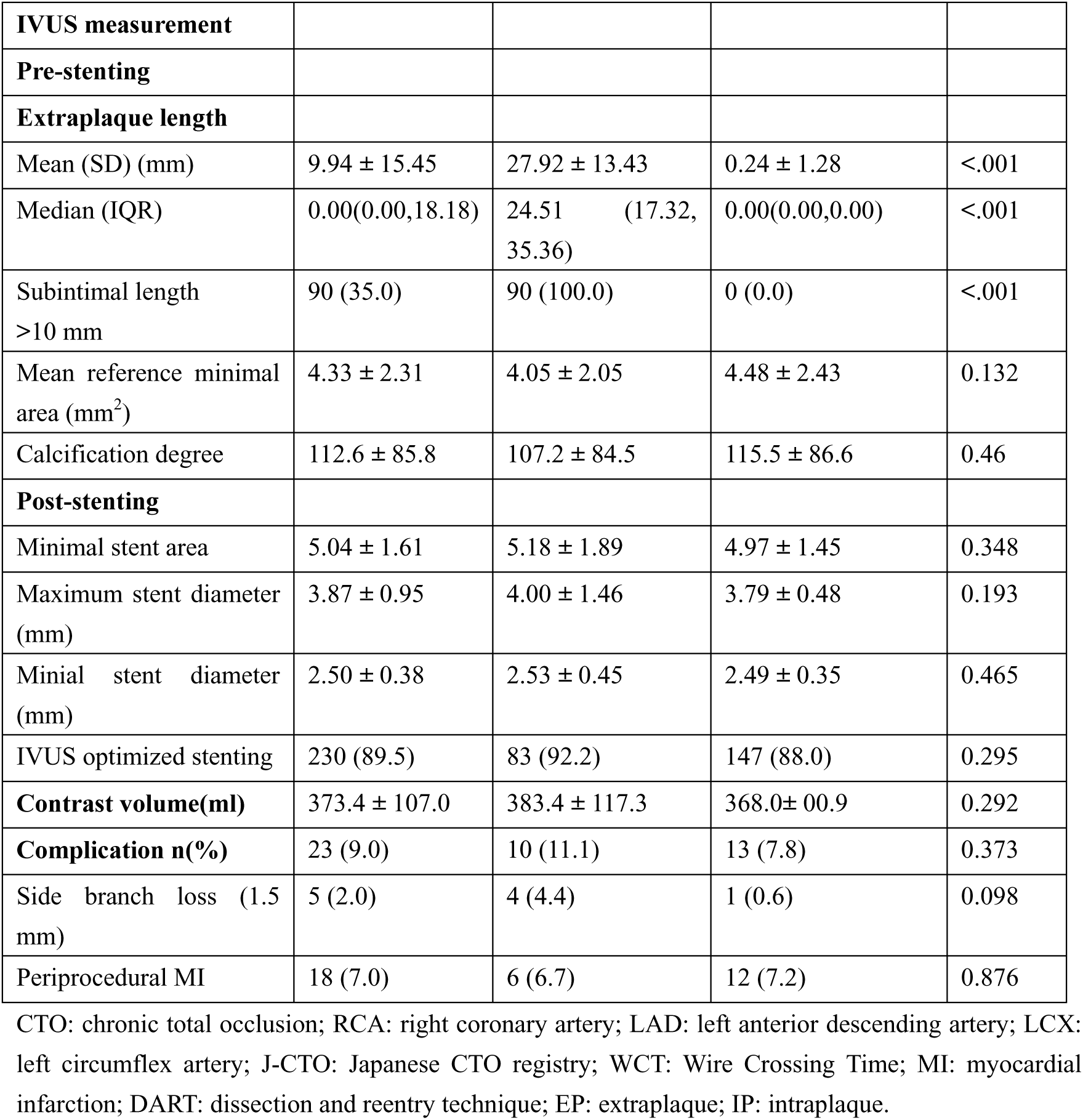
Baseline Lesion and Procedural Characteristics in EP and IP groups.

Among the 257 patients who underwent successful CTO PCI, 195 cases (76%) used AWE/RWE techniques, and 62 cases (24%) used the DART technique (Figure 2A). After IVUS examination, 20.5% (40/195) of the cases involved wire crossing in the subintimal space, and 9.7% (6/62) involved DART tracking in the intraplaque space. Finally, the discordance rate between the presumed and actual wire passage positions was 17.9% (Figure 2B). Combining the trial setup, specific lesions with SL >10 mm were assigned to the EP group; therefore, 6 out of 96 cases with subintimal tracking (3 cases in AWE, 2 cases in RWE, and 1 case in ADR) were assigned to the IP group, as shown in the trial flowchart in Figure 1.

**Figure 2.**
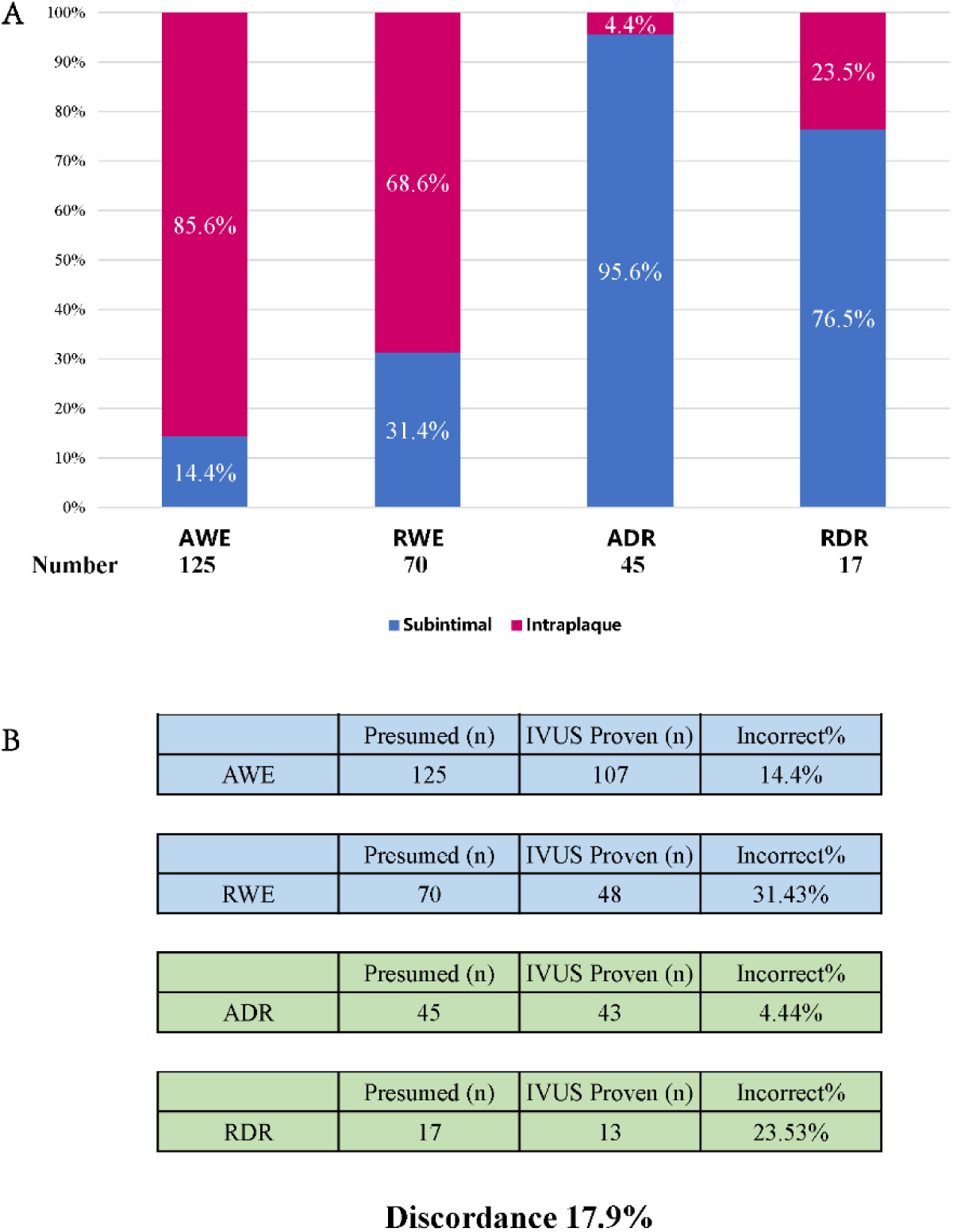
A: The proportion of four techniques in two groups; B: The discordance of wire position after IVUS assessment. AWE: Antegrade Wire Escalation; RWE: Retrograde Wire Escalation; ADR: Antegrade Dissection and Re-entry; RDR: Retrograde Dissection and Re-entry;

CTO lesions requiring the RDR technique were significantly more complex (mean J-CTO score: 2.82 ± 0.73) than those requiring WE strategies (2.16 ± 0.81 and 2.39 ± 0.82, respectively), or the ADR technique (2.24±0.88) (*p* < 0.05). The prevalence of guidewire tracking patterns among different J-CTO scores is shown in Figure 3.

**Figure 3:**
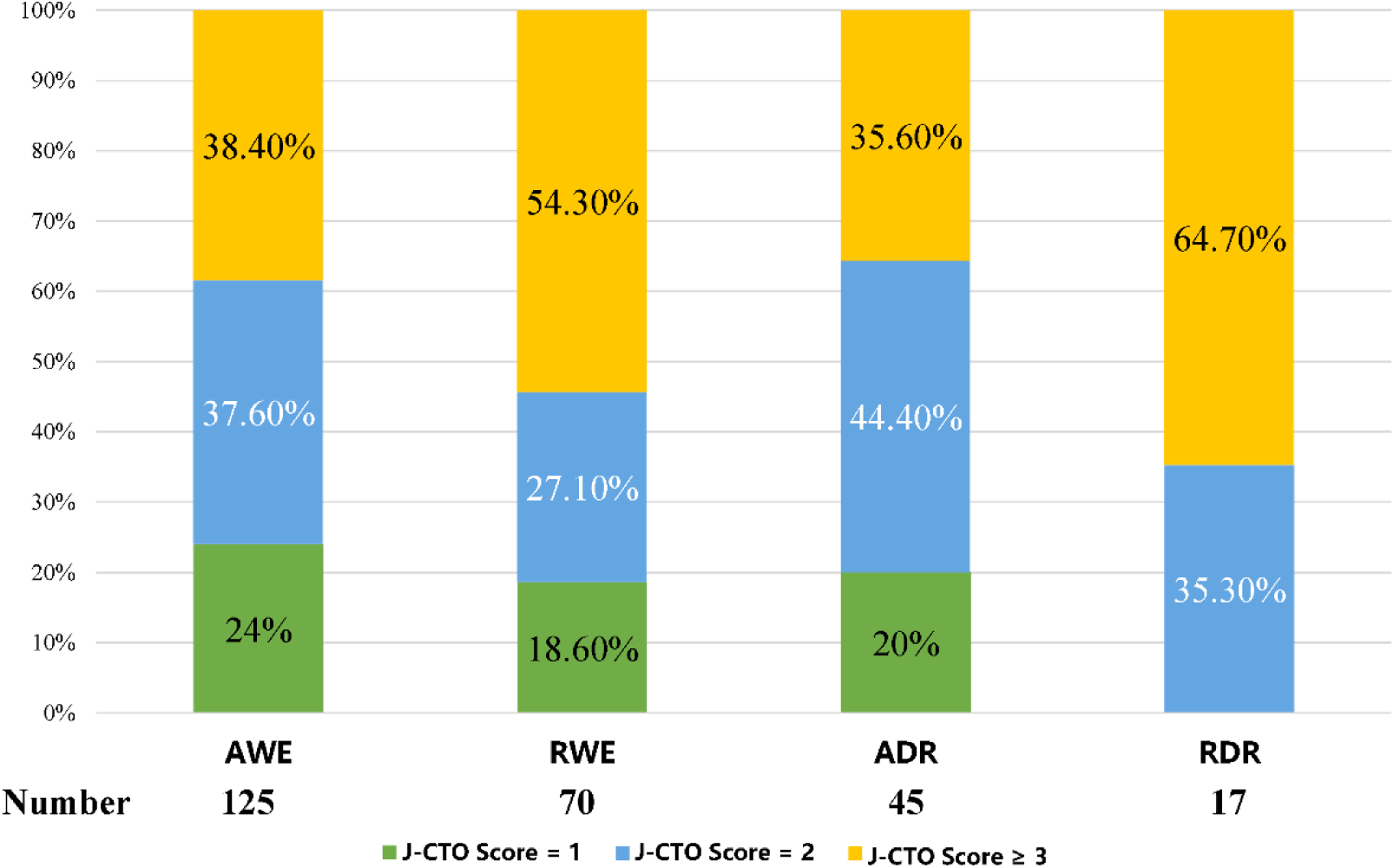
Proportion of J-CTO Score in Four Techniques. AWE: Antegrade Wire Escalation; RWE: Retrograde Wire Escalation; ADR: Antegrade Dissection and Re-entry; RDR: Retrograde Dissection and Re-entry; J-CTO: Japanese CTO registry.

### QCA and IVUS Results for the Procedure

QCA and IVUS results for both groups are presented in Table 2. QCA analysis showed that the mean occlusion length was 41.2 ± 15.7 mm in all cases, which was significantly longer in the EP group than in the IP group (46.6 ± 16.1 mm vs. 38.3 ± 14.8 mm, *p* < 0.001). The MLD was 2.66 ± 0.41 mm, and the minimal stent diameter (MSD) was 2.53 ± 0.45 mm, with no statistically significant difference between the two groups. The total stent length (TSL) was 70.0 ± 24.5 mm in all cases and was numerically longer in the EP group than in the IP group (73.8 ± 22.6 mm vs. 67.9 ± 25.3 mm, *p* = 0.066). The proportion of TSL > 50 mm was higher in the EP group than in the IP group (84.4% vs. 74.3%, *p* = 0.061), and showed a significant difference between the DART tracking group and the no-DART tracking group (88.7% vs. 74.4%, *p* = 0.018) (Table S2).

The median subintimal length measured by IVUS in the EP group was 24.5 mm (IQR: 17.3–35.4). 89.5% of the overall population underwent post-dilation after IVUS assessment, and the stent-related area and diameter values showed no significant differences between the two groups or between the DART tracking group and the no-DART tracking group.

The overall procedural complication rate was 9.0%. There was no statistically difference in the rate of side branch occlusion or peri-procedural MI between the EP and IP groups (*p* = 0.098 and *p* = 0.876) (see Table 2). The rate of side branch occlusion showed no significant difference between the DART tracking and no-DART tracking (4.4% vs. 0.6%, p = 0.098) (Table S2), and 3 out of 4 cases were caused by the ADR technique; however, there was no direct correlation between MI and side branch occlusion.

### QCA Results at the 13-Month Follow-up

The surveillance angiography was available for 70.8% (182/257) of all patients at the 13-month follow-up, and the median angiography time was 12.9 months (IQR: 12.1–15.4). The follow-up (FU) rate was 66.7% (60/90) in the EP group and 73.1% (122/167) in the IP group, with no significant difference (p = 0.26).

The QCA results for the same patients from the post-procedure and the 13-month FU are reported in Table 3. The occlusion length showed a significant difference between the EP and IP groups (46.5 ± 16.8 mm vs. 39.8 ± 15.4 mm; *p* = 0.007). However, no significant differences were observed between the two groups in terms of TSL and MLD post-procedure. At the 13-month angiographic follow-up, QCA results showed that the MLD was not significant difference between the two groups, nor were binary ISR or LLL. The rate of re-occlusion was lower (3.3% vs. 5.7%, *p* = 1.0). Additionally, the incidence of stent malapposition or aneurysm formation was very low in all patients (1.65%). Similar results were observed in both the DART and non-DART tracking groups (see Table S3).

**Table 3.** Quantitative Coronary Analysis at Post Procedure and Follow-up in EP and IP groups.

|  | **Total (n = 182)** | **EP (n = 60)** | **IP (n = 122)** | **P** |
| --- | --- | --- | --- | --- |
| **QCA measurement during PCI** |  |  |  |  |
| Occlusion length (mm) | 42.0 ± 16.1 | 46.5 ± 16.8 | 39.8 ± 15.4 | 0.007 |
| Total stent length (TSL, mm) | 69.7 ± 24.1 | 72.0 ± 21.7 | 68.6 ± 25.3 | 0.369 |
| Minimal lumen diameter (MLD, mm) | 2.65 ± 0.41 | 2.67 ± 0.45 | 2.63 ± 0.38 | 0.519 |
| Proximal- MLD (mm) | 3.06 ± 0.44 | 3.07 ± 0.46 | 3.05 ± 0.43 | 0.806 |
| Distal - MLD (mm) | 2.44 ± 0.42 | 2.48 ± 0.45 | 2.42 ± 0.41 | 0.378 |
| **QCA measurement at 13-mth follow-up** |  |  |  |  |
| MLD (mm) | 2.21 ± 0.65 | 2.23 ± 0.61 | 2.19 ± 0.67 | 0.701 |
| Proximal- MLD (mm) | 2.76 ± 0.57 | 2.72 ± 0.68 | 2.77 ± 0.51 | 0.565 |
| Distal - MLD (mm) | 1.92 ± 0.59 | 1.90 ± 0.60 | 1.94 ± 0.58 | 0.718 |
| Binary restenosis (in segment), n (%) | 19 (10.4) | 7 (11.7) | 12 (9.8) | 0.704 |
| LLL (mm) | 0.44 ± 0.63 | 0.44 ± 0.55 | 0.44 ± 0.67 | 0.962 |
| Reocclusion, n(%) | 9 (5.0) | 2 (3.3) | 7 (5.7) | 0.734 |
| Stent malposition/Aneurysm n(%) | 3 (1.7) | 2 (3.3) | 1 (0.8) | 0.253 |

IVUS: Intravascular ultrasound; QCA: quantitative coronary angiography; ISR: in-stent restenosis; LLL: late lumen loss; EP: extraplaque; IP: intraplaque.

### Clinical Outcomes at One-Year and Three-Year Follow-Up

A total of 257 individuals were included for a 1-year FU, with 90 cases in the EP group and 167 cases in the IP group. During the one-year FU, one patient from the IP group died of pulmonary hemorrhage. The clinical events are shown in Table 4 and Table S4. The overall MACE rate was 2.3%, with no significant difference between the two groups (*p* = 0.93) or between DART and no-DART groups (*p* = 0.163). The 3-year clinical FU showed that seven patients died. The clinical events are shown in Table 4 and Table S4. The total MACE rate was 11.7% (30/257), with no significant difference between the EP and IP groups (15.6% vs. 9.6%, *p* = 0.155). The TVR rate was 7.0% (18/257), with no significant difference between the two groups (10% vs. 5.4%, P=0.169), and the MACE rate was mainly driven by TVR. Similar trends were observed in both the DART and non-DART groups.

**Table 4.** MACE at Long-term Clinical Follow-up in in EP-s and IP-s groups.

| **MACE n (%)** | **Total (n = 257)** | **EP-s (n = 90)** | **IP-s (n = 167)** | **log-rank *P*** |
| --- | --- | --- | --- | --- |
| **MACE at 1 years** | 6 (2.3) | 2 (2.2) | 4 (2.4) | 0.930 |
| all-cause mortality | 1 (0.4) | 0 (0.0) | 1 (0.6) | 0.463 |
| cardiovascular mortality | 0 (0.0) | 0 (0.0) | 0 (0.0) | >0.999 |
| non-fatal MI | 0 (0.0) | 0 (0.0) | 0 (0.0) | >0.999 |
| TVR | 5 (1.9) | 2 (2.2) | 3 (1.8) | 0.82 |
| **MACE at 3 years** | 30 (11.7) | 14 (15.6) | 16 (9.6) | 0.155 |
| all-cause mortality | 7 (2.7) | 2 (2.2) | 5 (3.0) | 0.718 |
| cardiovascular mortality | 3 (1.2) | 1 (1.1) | 2 (1.2) | 0.955 |
| non-fatal MI | 5 (2.0) | 3 (3.3) | 2 (1.2) | 0.239 |
| TVR | 18 (7.0) | 9 (10.0) | 9 (5.4) | 0.169 |

- 5 MACE: major adverse cardiac events; MI: myocardial infarction; TVR: target vessel revascularization; EP: extraplaque; IP: intraplaque.

The Kaplan–Meier plot showed the TVR cumulative incidence rate at the 3-year FU, with no statistical difference between the two groups (p = 0.11). A similar trend was observed in DART and no-DART tracking (*p* = 0.83) (Figure S4-1). The four types of techniques did not have a significant impact on the TVR rate (*p* = 0.69) (Figure S4-2).

### Univariate and Multivariable Analyses of ISR and MACE Rates

These values in terms of wire crossing time, LCX-CTO, and TSL were found to be independent variables associated with ISR occurrence in univariate analysis. Multivariate analysis showed that wire crossing time of <30 minutes (OR = 0.16, 95% CI: 0.03–0.78, *p* = 0.024) was a protective factor, while LCX-CTO (OR = 10.49, 95% CI: 1.99–55.24, *p* = 0.006) and TSL of ≥50 mm (OR = 6.68, 95% CI: 1.04–42.76, *p* = 0.045) were independently associated with the occurrence of ISR (see Figure 5).

**Figure 4.**
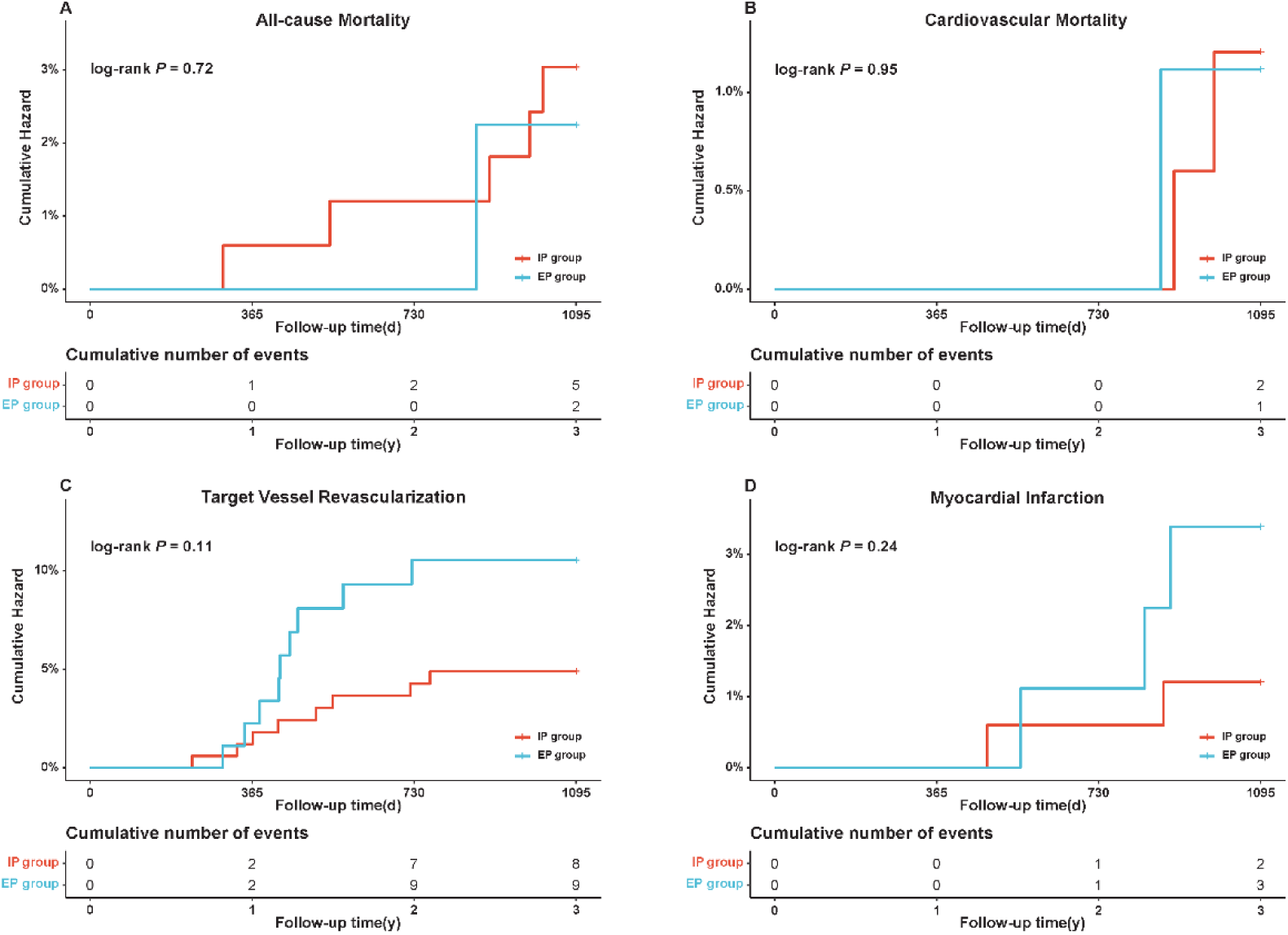
Cumulative Distribution Functions curves for LLL and ISR at 13-monthangiographic FU in EP and IP groups. (A) cumulative distribution functions curve for LLL; (B) cumulative distribution functions curve for ISR; LLL: Late Lumen Loss; ISR: In-Stent Restenosis; Follow-Up: FU; EP: Extra-plaque; IP: Intraplaque

**Figure 5.**
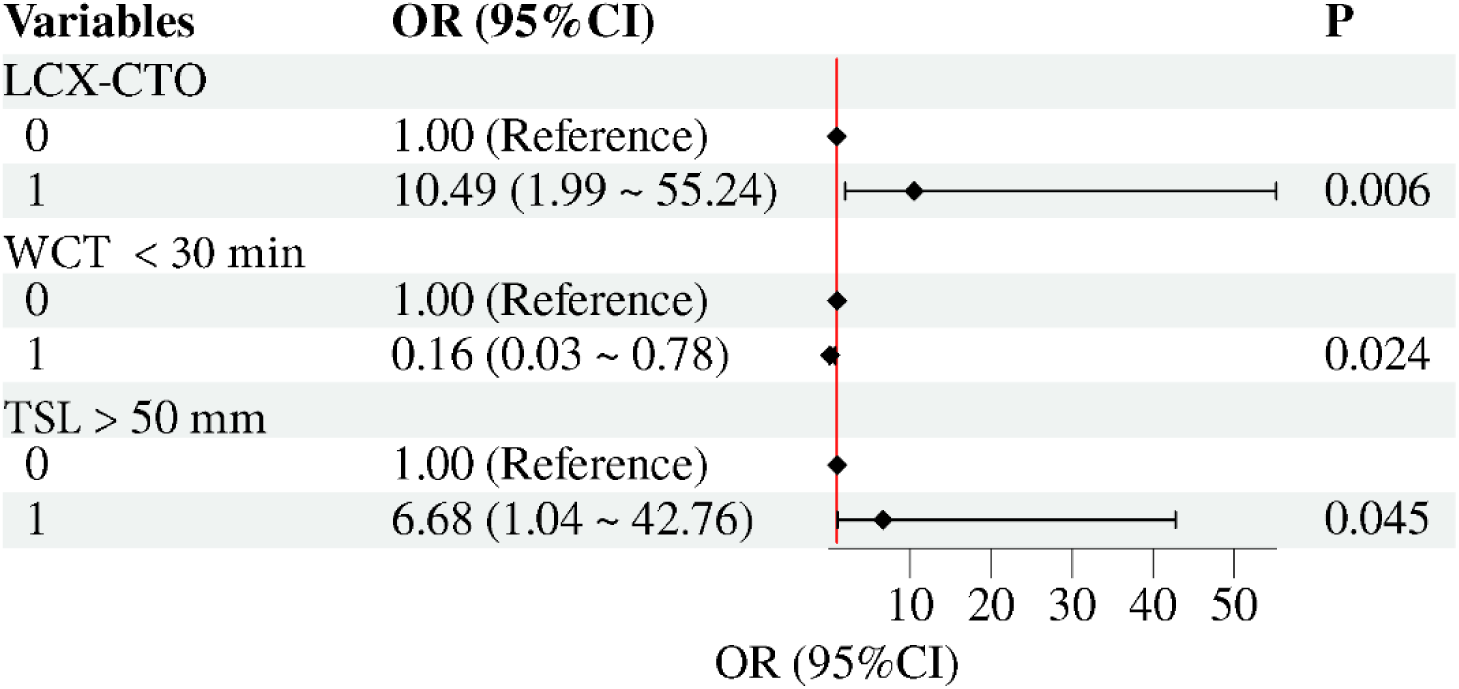
Cumulative Incidence Curves for 3-yr MACE clinical FU in EP and IP groups. (A) cumulative hazard for all-cause mortality; (B) cumulative hazard for cardiovascular mortality; (C) cumulative hazard for TVR; (D) cumulative hazard for MI; MACE: Major Adverse Cardiac Events EP: Extra-plaque; IP: Intraplaque; TVR: Target Vessel Revascularization (TVR); MI: Myocardial Infarction

As shown in Figure 6, the results of multivariate analysis for 3-year MACE. Age >70 years was independently correlated with all-cause mortality (log-rank *p* = 0.004). Wire crossing time of >2 hours was associated with the occurrence of TVR (*p* = 0.010), and multivessel disease was associated with the occurrence of myocardial infarction (*p* = 0.013).

**Figure 6.**
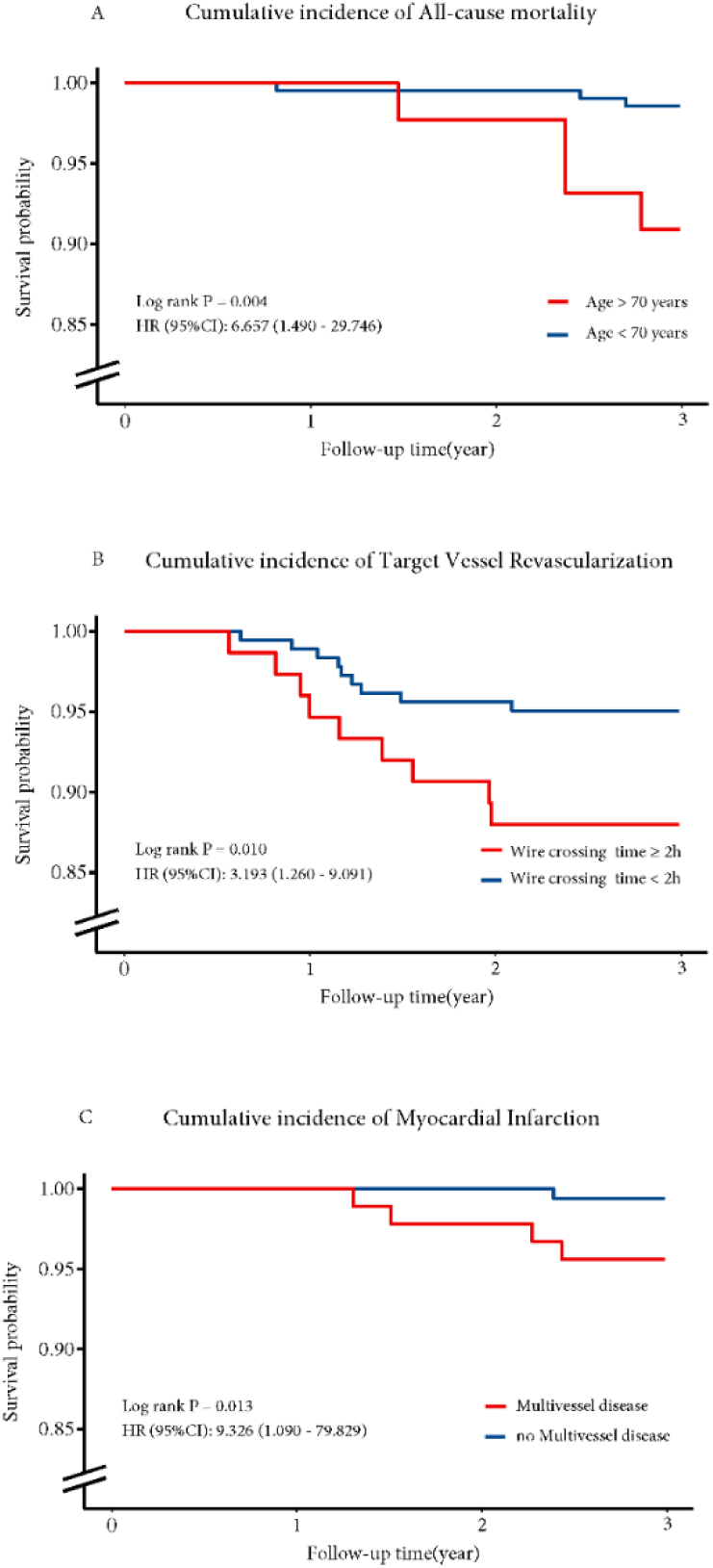
Cox Regression for cumulative incidence of 3-yr MACE FU. A: Cumulative Incidence of all-cause modality at 3-yr FU B: Cumulative Incidence of target vessels revascularization at 3-yr FU C: Cumulative Incidence of myocardial infarction at 3-yr FU HR: Hazard rate; CI: Confidence Interval

## DISCUSSION

Our trial represents one of the largest prospective studies comparing the long-term clinical outcomes between extraplaque and intraplaque tracking following contemporary CTO PCI recanalization techniques. These are the main findings of the study: (1) Four tracking techniques were required for successful recanalization in the contemporary CTO PCI procedure; (2) The intended crossing technique does not correspond with the actual crossing strategy in approximately 21% of WE cases and 10% of DART cases; (3) The angiographic outcomes and long-term clinical outcomes were similar between EP and IP tracking, regardless of the CTO crossing strategy; (4) Total stent length >50 mm and LCx CTO were independently associated with ISR, and long wire crossing time of >2 hours was associated with a higher TVR rate.

### Disease Complexity and Unique IVUS Assessment

The enrollment of patients and angiographic follow-up in our trial were affected by the COVID-19 epidemic in China. Ultimately, we completed 83.3% (257) of the originally planned 300 patients. The CTO lesions were more complex in all patients; 95.7% of CTO lesions had a length ≥ 20 mm, which was higher than that reported in three existing studies, which had reported rates of about 40–50% [20–22]. Meanwhile, the mean lesion length was 41.2 ± 15.5 mm, which was longer than the mean lesion length reported in a comparable CTO study (36.3 ± 22.0 mm [17]) and in another study (37.43 ± 26.67 mm [12]). In our study, 79.8% of patients had a J-CTO score of ≥2, and 44.0% had a J-CTO score of ≥3, which were consistent with the higher J-CTO scores reported by Wilson WM [14] and in the ISAR-OCT-CTO study [15].

To our knowledge, five large studies [11, 16–17, 21–22] have utilized IVUS to evaluate wire position after different tracking techniques. These studies have shown that assessing guidewire courses based on technique resulted in classification bias, particularly in cases of WE tracking. In a meta-analysis [23], the wire position was different from expected in approximately 10% of antegrade cases and 15%–30% of retrograde cases. In the Consistent CTO study [17], the discordance rate was 15.8%. In our study, using the DART tracking technique, the discordance rate was 17.9%, indicating that predicting the wire escalation course was more challenging than predicting the DART wire course. Another concern is that only 2 studies have measured the subintimal length [21, 22]. In these two studies, more than 50% of the overall cases involved subintimal tracking, and one of them reported that 68% of cases had a subintimal length > 10 mm (median 21.5 mm, IQR 7.3–37.2). However, no study has addressed that a short subintimal track could have different clinical implications compared to a long one. In this prospective study, we cautiously explored the long-term clinical outcomes in patients with a specific subintimal length greater than 10 mm. Among the cases without DART tracking, 18% (35 out of 195) had a subintimal length > 10 mm, which highlights the importance of IVUS assessment.

### Angiographic and Long-term Clinical Follow-up Outcomes

The clinical ischemia symptoms caused by TLR/re-occlusion in patients are not obvious due to the resupply flow by collateral vessels [25]. Therefore, surveillance angiography is usually scheduled to reassess distal vessel run-off and remodeling, and to avoid underestimating the occurrence of restenosis or re-occlusion. Only 5 studies [11, 15–18] among those investigating the clinical outcomes of intraplaque and subintimal stenting included systematic angiographic FU. In our trial, according to angiography analysis, the EP group showed a re-occlusion rate of 5.6% and a segment restenosis rate of 11.1%, comparable to the rates reported in the consistent CTO study (3.4% and 14.5%, respectively) [17].

The angiographic parameters showed no significant differences, as reported in other angiographic follow-up studies [15–16, 18]. Our results confirm the sustained efficacy of subintimal stenting using the contemporary DART technique. Studies have reported that IVUS-guided stenting of CTO lesions is associated with less LLL and a lower incidence of stent restenosis [26]. In our study, up to 89.5% of all cases underwent post-dilation after IVUS assessment to achieve the optimal stent area.

Despite the complexity of the recruited cohort, the incidence of adverse events during the one-year FU was low, with 2.2% for both TVR and MACE rates in the EP group. Moreover, this TVR rate was lower than the previously reported rates of 4.5–7.0% in earlier CTO cohorts using the DART technique [13,14]. At the 3-year FU, 10.0% of TVR rate and 15.6% of MACE rate in the EP group, which were comparable to those in the consistent CTO study reporting 11.9% TVR and 17% MACE rate at the 24-month FU [17]. The EP group and the DART tracking technique did not increase the TVR rate or the occurrence of MACE, which was similar to many studies on DART tracking in clinical outcomes [17,18]. These results demonstrated that long subintimal stenting was durable, regardless of the tracking technique used, in more complex CTO lesions.

### Predictors of ISR and MACE Rates

The multivariate analysis for ISR showed that LCx-CTO was an independent predictor, likely related to the greater tortuosity of the LCx vessel and the insufficiency of interventional collaterals [27]. Another important finding was the correlation between TSL > 50 mm and ISR occurrence. Due to the longer occlusion lesion length (41.2 ± 15.7 mm) in the present data, it was anticipated that longer stent lengths would be required to treat these more complex lesions, as reported in previous CTO cohort studies [17, 20–22]. This is likely to increase the risk of restenosis [21, 22] and TLR; however, there is no direct relationship with subintimal wire tracking [20]. In the present study, DART tracking increased the proportion of TSL > 50 mm compared to no-DART tracking, which further suggests that selecting appropriate CTO lesions and applying DART tracking by experienced CTO experts can help minimize the subintimal length.

The multivariate analysis for MACE occurrence showed that wire crossing time of >2 hours was an independent predictor. The long wire crossing time practically indicates the complexity of CTO lesions, corresponds to a higher J-CTO score, and suggests that more complex techniques and invasive devices are used, which may lead to more complications [28]. Until now, few studies have reported prolonged wire crossing time as contributing to an increase in TVR, and that further research is needed to investigate this association. Meanwhile, we found that multivessel lesions with CTOs were independently associated with the occurrence of MI. Previous studies have shown that the strongest predictors of incomplete revascularization are the number of diseased vessels and the presence of CTOs [29].

Therefore, we demonstrated that no matter the subintimal length or tracking technique used during the treatment of very complex CTO lesions, favorable angiographic and clinical short- and long-term outcomes can be achieved, and contemporary CTO PCI techniques can be safely applied when indicated.

### Limitations

Some limitations should be considered when interpreting the results of the present report. First, due to the impact of the COVID-19 pandemic, only 257 patients were enrolled, which was fewer than the originally planned 300 cases, and the angiographic follow-up rate was relatively low. Second, we included only patients who underwent successful CTO PCI because our study aimed to evaluate the clinical outcomes of extraplaque tracking by different crossing techniques rather than procedural outcomes. Third, QCA and IVUS analyses were not conducted in an independent core laboratory. Finally, although this was a multicenter registry with several operators involved, our study findings might not be generalizable to other institutions or interventionalists who are not experienced with the hybrid algorithm.

### Conclusions

The specific long extraplaque tracking during CTO PCI procedures might have comparable angiographic and long-term clinical outcomes to intraplaque tracking. This finding supports the broader application of contemporary CTO recanalization techniques, particularly when performed by experienced CTO operators to minimize subintimal length.

## DECLARATION

### Data Availability

The data that support the findings of this study are available from the corresponding author upon reasonable request. All such requests should be submitted to Prof. Li Chengxiang.

### Ethical Approval

The present study was approved by the Institutional Review Committee of the First Affiliated Hospital of Air Force Military Medical University and was conducted in accordance with the Declaration of Helsinki. All patients provided written informed consent for the use of their clinical data in clinical research.

